# Prevalence and Associated Factors of Diabetic ketoacidosis among Patients Living with Type 1 Diabetes in Makkah Al-Mukarramah City

**DOI:** 10.1101/2021.10.28.21264912

**Authors:** Raghad Alhajaji, Khalid Almasodi, Afaf Alhajaji, Ahmad Alturkstani, Mayada Samkari

## Abstract

**Objective:** To assess magnitude of diabetic ketoacidosis (DKA) among type-1 diabetics and to identify associated risk factors.

**Methods:** A cross-sectional study was conducted among 236 type-1 diabetics in Makkah Al-Mukarramah City, Saudi Arabia.

**Results:** Among participants, 59.3% were males, 44.1% were diabetic for more than 5 years, while 70.8% reported past history of DKA. The main causes of DKA were “first presentation of the disease” (40.9%), and “discontinued treatment” (37%). The HbA1c among 53.6% was above 9%. Almost all cases who experienced DKA were hospitalised (98.8%). Out of them, 9 (5.4%) suffered complications. Female patients were more likely to suffer from episodes of DKA than males (76% and 68.3%, respectively). Most patients whose parents’ highest education was primary level had DKA more frequently than those whose parents’ had postgraduate education. Patients with unemployed fathers had significantly higher frequency of DKA (p=0.004). Ketoacidosis was significantly more frequent among patients with parents’ consanguinity (p<0.001). Patients who had their current HbA1c level exceeding 9% had positive history of DKA compared to those with HbA1c level ≤7% (87.9% and 28.6%, respectively, p<0.001).

**Conclusion:** Most type-1 diabetics experience DKA, mainly with their first presentation of disease or due to discontinuation of treatment. DKA tends to occur more frequently among female patients, those with less educated parents or when their parents are relatives.

**ADVANCES IN KNOWLEDGE:**

1. This study rang a warning bell towards parents’ consanguinity among type-1 diabetics as a risk factor for diabetic ketoacidosis.
2. The current search revealed the lack of public’s awareness about type-1 diabetes and its complications as well as their low compliance towards its treatment, since diabetic ketoacidosis occurred mainly at the first presentation of the disease among diabetics, or due to treatment discontinuation.

**APPLICATION TO PATIENT CARE:**

1. This study indicated the importance of raising the public’s awareness regarding early symptoms of diabetes among their children so as to be ready to seek medical advice as early as possible before the occurrence of complications.
2. Health education messages should be repeatedly broadcast via mass media explaining the hazards associated with consanguineous marriages.
3. Health care providers should stress to diabetic patients and/or their caregivers the importance of compliance to treatment.

## INTRODUCTION

During childhood and adolescence, type-1 diabetes mellitus (T1DM) is considered the most common endocrine-metabolic disorder and one of the major threats to human health ^(1-2)^. Almost one in 300 youths develop T1DM (3). Worldwide, it has been reported that the incidence of T1DM is increasing by 3-4% per year ^(4)^.

Diabetic ketoacidosis (DKA) usually occurs as a result of insulin deficiency. It is a serious acute complication of diabetes mellitus (DM), which accounts for most hospitalizations due to severe insulin deficiency ^(5)^. It consists of the biochemical triad of ketonaemia, hyperglycaemia and acidaemia ^(6)^. Among children, the criteria for diagnosis of DKA includes blood glucose above 11 mmol/L, venous pH less than 7.3, or bicarbonate less than 15 mmol/L, and ketonaemia with ketonuria ^(7)^.

Although major advances have been achieved in the fields of care for diabetic patients, DKA continues as a significant cause of morbidity and mortality ^(8)^. It is frequently the main presenting symptom for new-onset cases in 25% to 30% of T1DM cases ^(9)^.

The incidence of DKA is difficult to establish, but it continues to increase, accounting for about 140,000 hospitalizations in the US in 2009 and more than 500,000 hospital days annually ^(10)^. Even with the promising statistics and raised awareness, the occurrence of DKA continues to be as high as 30% in children with T1DM ^(11)^.

The clinical presentation of DKA usually develops rapidly, over a period of less than 24 hours. Several days before development of DKA, several symptoms may develop, i.e., polyuria, polydipsia, and weight loss. The presenting symptoms usually include vomiting and abdominal pain ^(12)^. Physical examination of a patient with DKA shows signs of dehydration, e.g., loss of skin turgor, dry mucous membranes, tachycardia, and hypotension. Patient’s level of consciousness varies from being full alert to loss of consciousness ^(13)^.

Although the diagnosis of DKA can be suspected on clinical grounds, confirmation is usually based on results of laboratory tests. The most widely used diagnostic criteria for DKA in the past was a blood glucose level more than 250 mg/dL, a moderate degree of ketonaemia, serum bicarbonate less than15 mEq/l, arterial pH less than 7.3, and an increased anion gap metabolic acidosis ^(13)^.

It is possible to prevent DKA by the establishment of better access to medical care, proper health education, and ensuring effective communication with health care providers during an intercurrent illness. It is also essential that family members become involved. Therefore, they should be educated on insulin regimen and patient’s blood glucose assessment. Moreover, a written care plan should be provided to diabetic patients and/or their caregivers, as this is essential to enhance their understanding of the importance of diabetes self-management ^(14)^.

The use of ketone-meters that detect blood β-hydroxybutyrate has also been shown to help early detection and management of ketosis, which may decrease the need for specialized care. Short-acting insulin may be administered with fluids, early on for the prevention of DKA (15).

The incidence of T1DM in Saudi Arabia total number of cases of T1DM in children under the age of 12 years was 22 with an estimated prevalence of 106.7/100,000 (^16)^. The incidence rate of T1DM is growing in Saudi Arabia(^17)^.

DKA is the most severe health problem among diabetic children and adolescents ^(5)^. It is typically caused by treatment non-compliance, i.e., shortage of insulin and may be precipitated by several factors, e.g., infections. Although DKA can be a life-threatening event for type-1 diabetics, it is a preventable condition. Recent advances in diabetes management could not minimize prevalence of DKA among children with T1DM ^(11)^. Despite the severity of DKA, research examining the event is limited in the empirical literature. Therefore, the identification of prevalence of DKA and its associated risk factors is a pressing necessity ^(18)^.

This study aimed to assess prevalence of DKA and to identify risk factors associated with it among Saudis with T1DM in Makkah Al-Mukarramah City.

## METHODOLOGY

A cross-sectional study design was followed at the Diabetes and Endocrine Unit in the Maternity and Children’s Hospital, and the Diabetes and Endocrine Center in Herra General hospital in the Holy City of Makkah Al-Mukarramah, Saudi Arabia. This study received the approval of the Ethical Research Committee of Makkah Al-Mukarramah Region on May 31^st^, 2018. The study was conducted between January 2018 and July 2018 and included 236 Saudi type-1 diabetic patients aged 1-19 years.

Based on relevant literature, a study questionnaire was designed in simple Arabic Language by the researcher. It comprised the following:

- Personal data: Age, gender, duration of diabetes, parents’ education, parents’ employment status, consanguinity between parents and family history of diabetes.
- DKA data: Number of DKA incidents, expected cause(s) for DKA, hospitalization, complications, receiving health education at the Diabetes Clinic.
- Laboratory findings: HbA1c level.

Parents’ consanguinity was classified according to Rohde et al. (^19)^, as follows:

- First degree consanguinity: If parents share grandparents but have different parents (i.e., first degree cousins);
- Second degree consanguinity: If parents share great grandparents but have different grandparents (i.e., second degree cousins).

The study questionnaire validity (face and content) was assessed by three academic professors of Community Medicine.

A pilot was conducted on 22 diabetics, aiming to test the clarity and wording of the study questionnaire. Moreover, test-retest reliability of responses for included statements was assessed by applying the study questionnaires twice to the same participants, one week apart and the correlation coefficients for each response was calculated. Moreover, internal consistency of study questionnaire was assessed by applying the Cronbach’s alpha coefficient.

The study settings were visited by the researcher during June and July 2018. All type-1 diabetic patients attending the Endocrine Clinic (and their caregivers) were briefed regarding the objectives of the study and were then invited to participate in the study. During data collection, participants were consecutively included in the study. The researcher then distributed the self-administered questionnaire sheet to each participant (or his/her caregiver). The questionnaire sheets were then collected immediately after being filled. The researcher repeated the daily visits till the required sample size was fulfilled.

The Statistical Package for Social Sciences (SPSS), version 25, was used for data entry and statistical analysis. Descriptive statistics (e.g., number, percentage, mean, range, standard deviation) and inferential statistics, using chi-square “χ^2^” test was applied. P-values <0.05 were considered as “statistically significant”.

All necessary official permissions were secured by the researcher before the start of the data collection. Before the data collection, all patients and their caregivers were verbally informed about the study objectives, and a written form (informed consent) was fulfilled. Confidentiality and privacy were fully secured for all patients.

## RESULTS

### Characteristics of the study group

Most patients were males (59.3%), their mean age was 10.7±4.3 years. Regarding parental characteristics, Table 1 demonstrates that almost two thirds of the fathers had either secondary level of education (34.9%) or university qualifications (32.1%) in addition to 7.6% who had postgraduate degrees, with comparable percentages in mothers where 28.1% had secondary level of education and 35.5% had university qualifications and 2.3% had postgraduate degrees. While the majority of fathers (66.8%) had jobs; only 27.3% of the mothers indicated that they had jobs. Consanguinity was identified in almost one half of the parents (51.1%), out of them 34.9% shared the same grandparents.

**Table 1:**
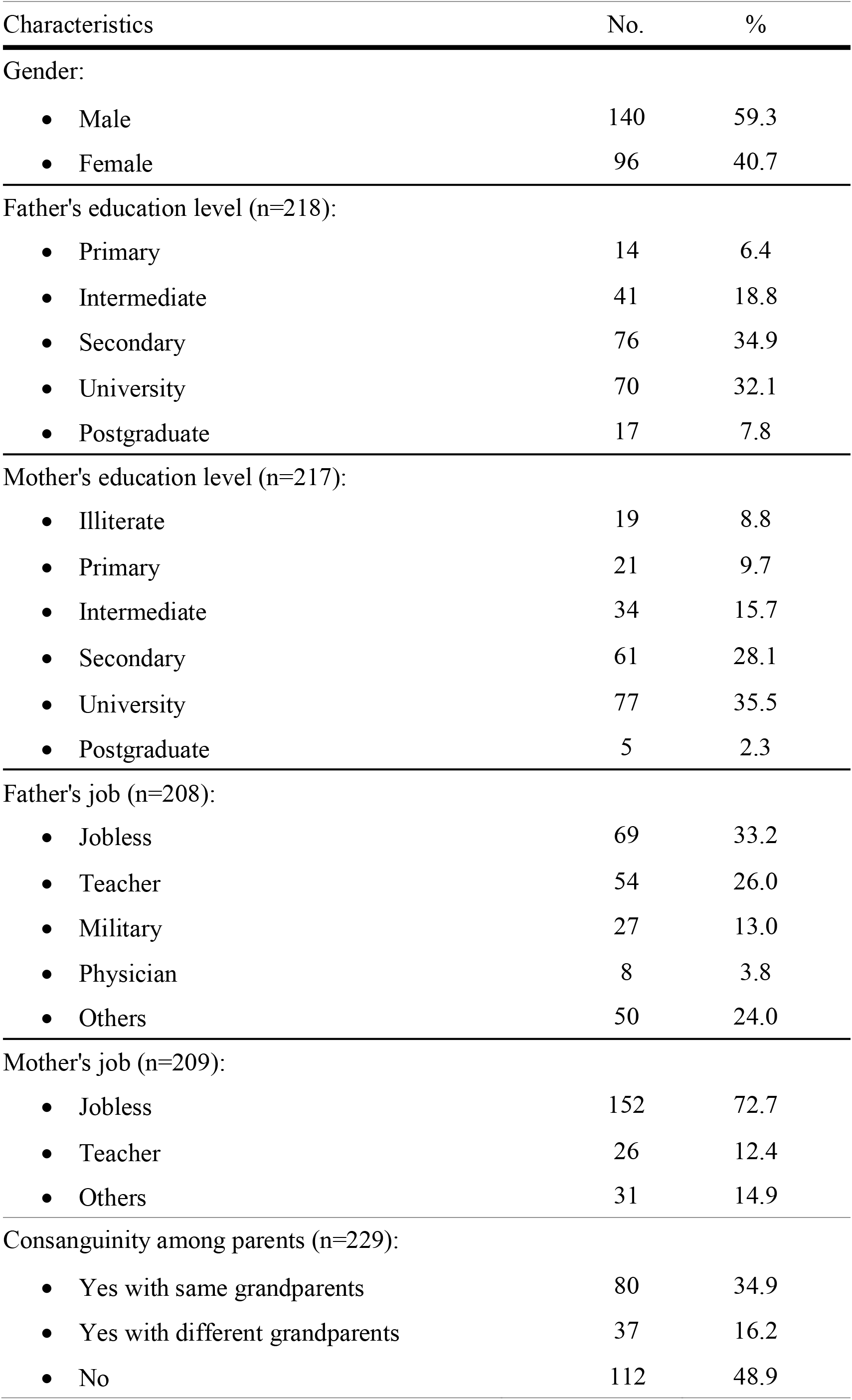
Demographic characteristics of the patients (n=236)

Regarding clinical characteristics of the patients, Table 2 shows that most patients (84.5%) had been diagnosed with diabetes mellitus more than one year earlier. Almost one half of the patients (48.3%) had positive family history of diabetes mellitus; out of them, 12.7% in first degree and 4.7% in both first and second degree relatives. Respecting the last reading of HbA1c level, it was found that the overwhelming majority of the patients (91%) had HbA1c level exceeding 7%, out of whom, there was 53.6% who had HbA1c level more than 9%.

**Table 2:**
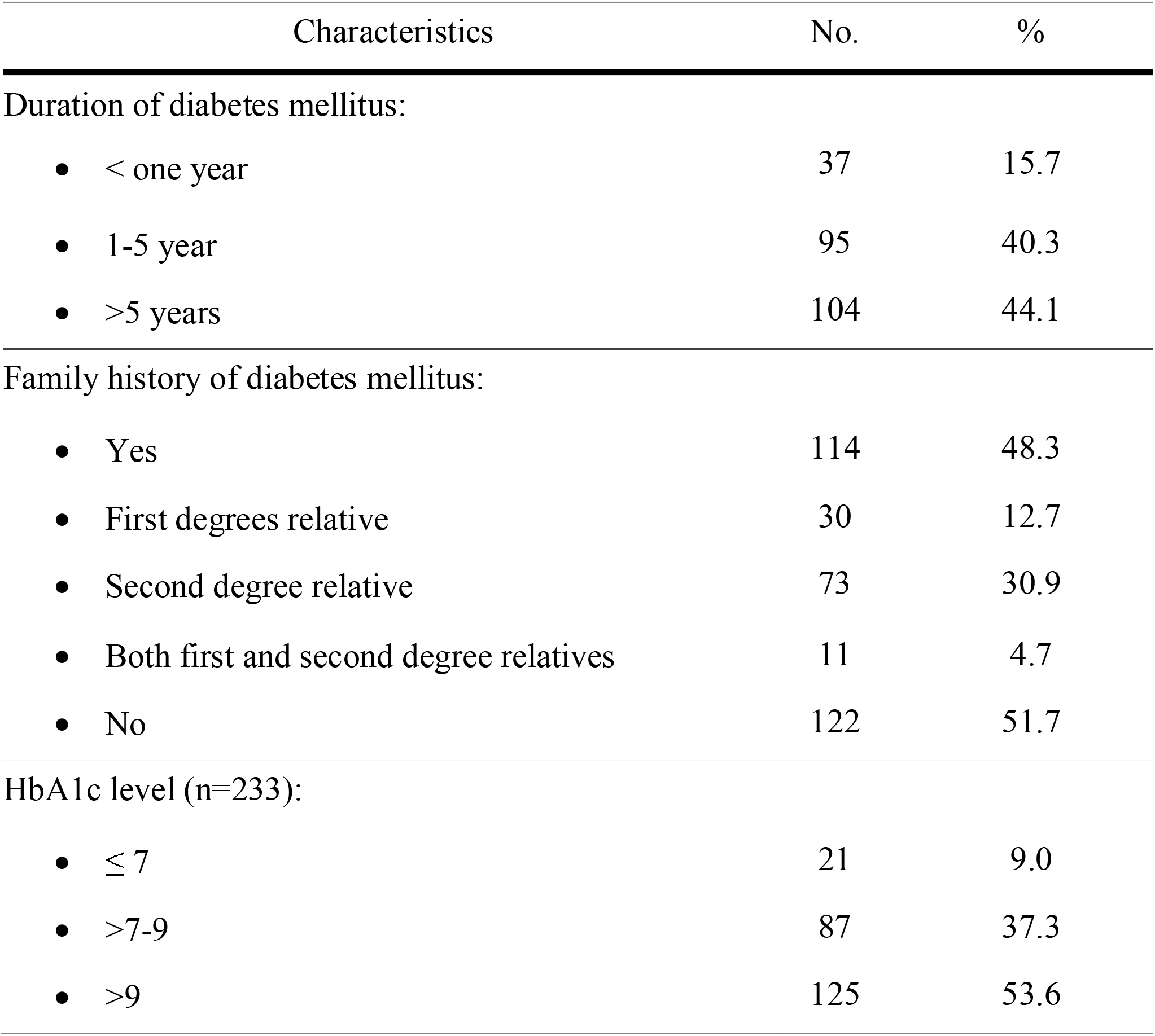
Clinical characteristics of the patients (n=236)

### History of Ketoacidosis

Figure 1 shows that most patients (70.8%) experienced ketoacidosis before. Out of them, there were 30.9% who had it once, 17.4% had it twice, and a total of 22.3% who had three or more episodes of ketoacidosis before.

**Figure (1):**
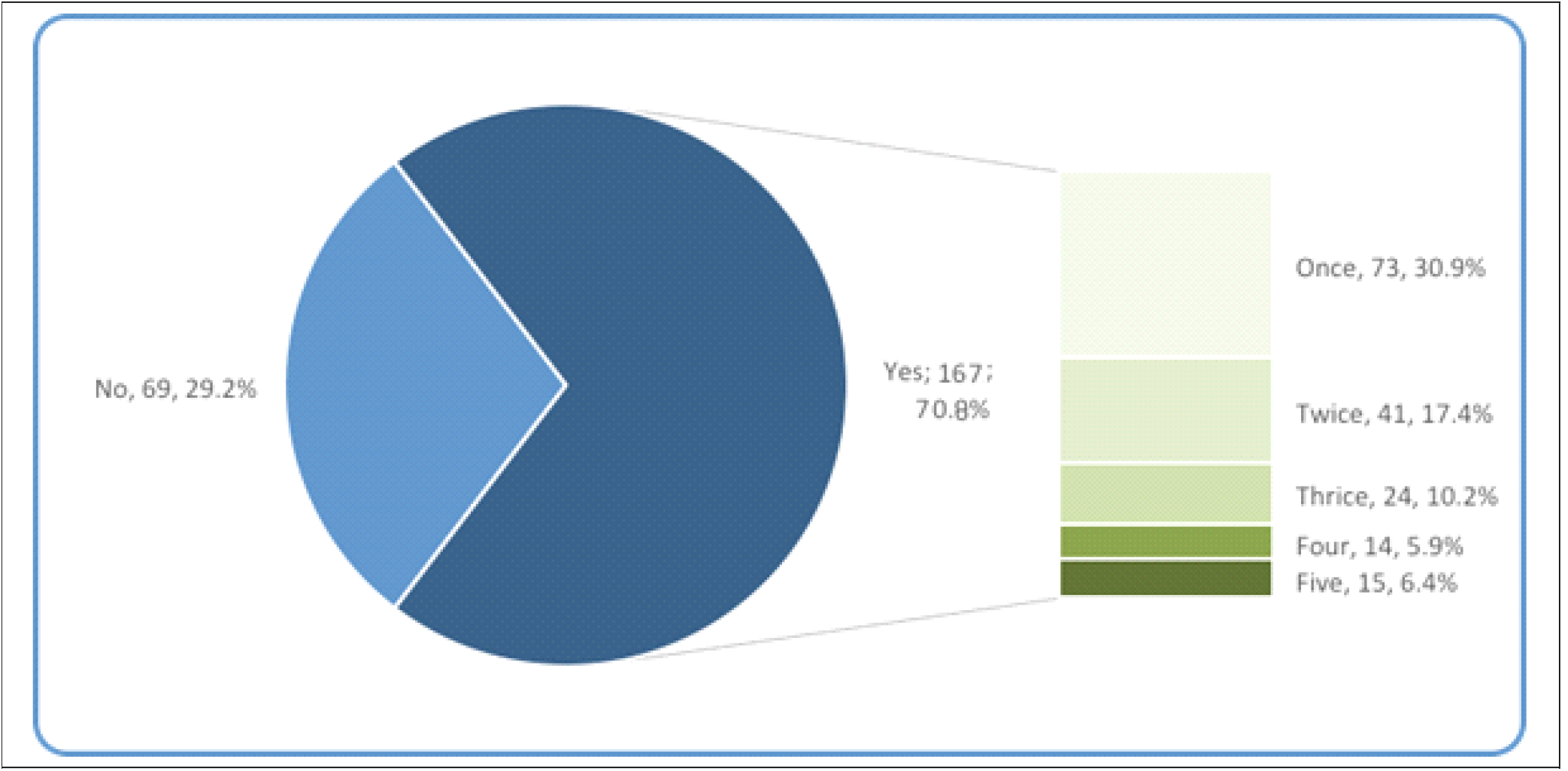
History of ketoacidosis among T1DM patients

Figure 2 shows that among 167 patients who experienced ketoacidosis before, for 41.9% it was their first presentation of diabetes mellitus. In 37.7%, ketoacidosis was attributed to discontinuation of treatment, and in 10.8%, it was attributed to non-adherence to diet.

**Figure (2):**
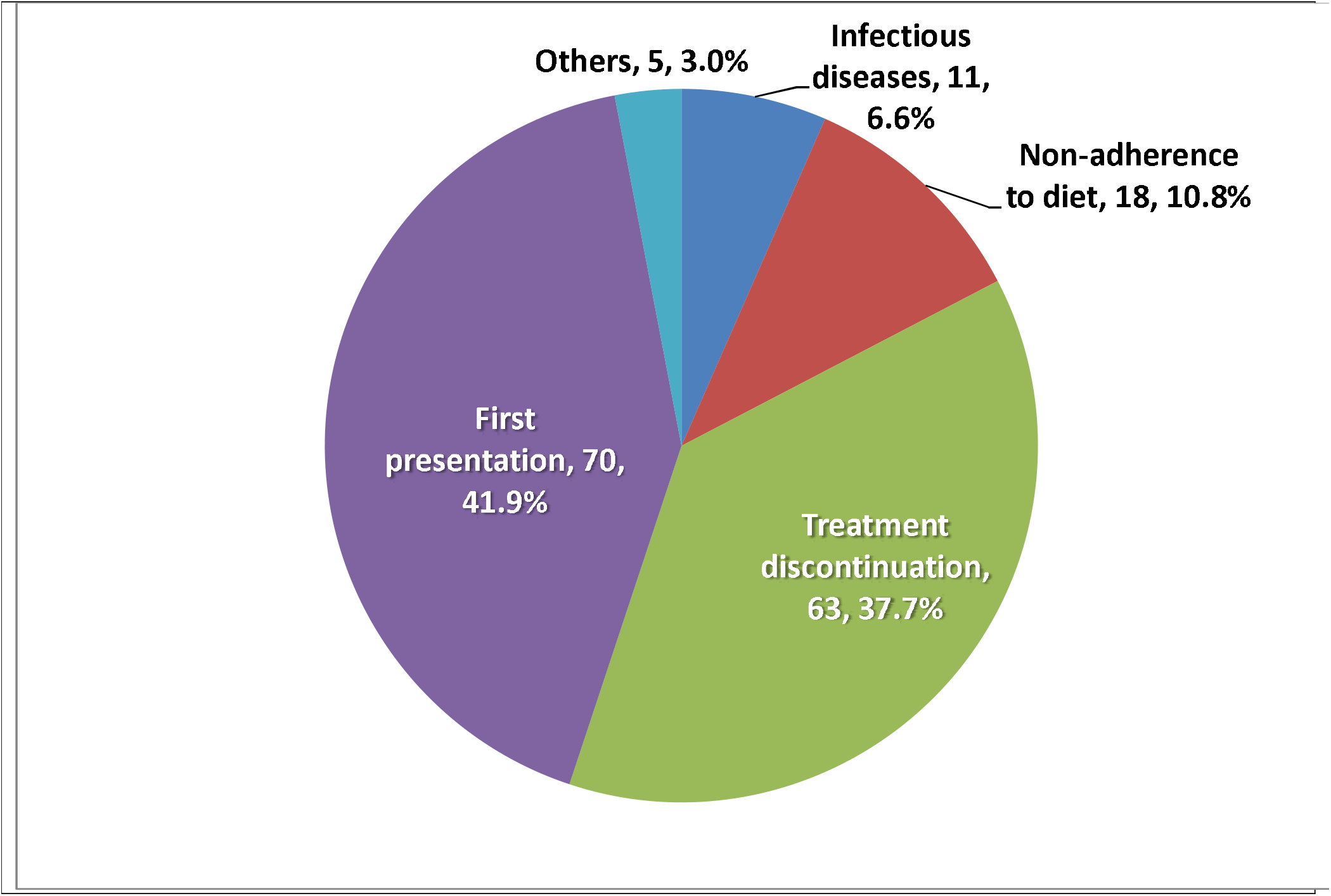
Predisposing factors of ketoacidosis among T1DM patients (n=167).

It is to be noted that almost all cases who experienced DKA were admitted to hospitals (165; 98.8%). Out of them, only 9 (5.4%) suffered from complications.

### Factors associated with ketoacidosis

Table 3 demonstrates that, although female patients were more likely to suffer from episodes of DKA than males (76%, 68.3%, respectively), this difference is not statistically significant. Also, despite the apparent difference in the frequency of DKA according to education levels of the fathers, where 92.9% of the patients whose fathers had primary level of education had history of ketoacidosis compared to only 52.9% of patients whose fathers had postgraduate degrees, this difference was not statistically significant. The same was also observed with regards to mothers’ educational levels, where the highest frequency was recorded in patients with illiterate mothers (89.5%), while the lowest was observed in patients whose mothers were university qualified (59.7%). Moreover, no statistically significant difference was observed according to mothers’ jobs.

**Table 3:** Characteristics of ketoacidosis among type 1 diabetics

| Characteristics | No. | % |
| --- | --- | --- |
| History of ketoacidosis |  |  |
| • Absent | 69 | 29.2 |
| • Present | 167 | 70.8 |
| Frequency of ketoacidosis |  |  |
| • Once | 73 | 30.9 |
| • Twice | 41 | 17.4 |
| • Three times | 24 | 10.2 |
| • Four times | 14 | 5.9 |
| • Five times | 15 | 6.4 |
| Predisposing factors for ketoacidosis |  |  |
| • First presentation | 70 | 41.9 |
| • Treatment discontinuation | 63 | 37.7 |
| • Non-adherence to diet | 18 | 10.8 |
| • Infectious diseases | 11 | 6.6 |
| • Others | 5 | 3.0 |

Frequency of DKA was significantly higher among patients with jobless fathers (79.7%), compared to patients whose fathers were employed (p-0.004). Moreover, ketoacidosis was significantly higher in presence of consanguinity of the parents, whether with same grandparents (86.5%) or with different grandparents (86.2%), compared to only 55% where there is no consanguinity (p<0.001).

As shown in Figure 3, **there was** no significant difference in age of those who had history of DKA and those who did not have DKA (Mean±SD: 10.5±4.6 and 11.1±3.6, respectively).

**Figure (3):**
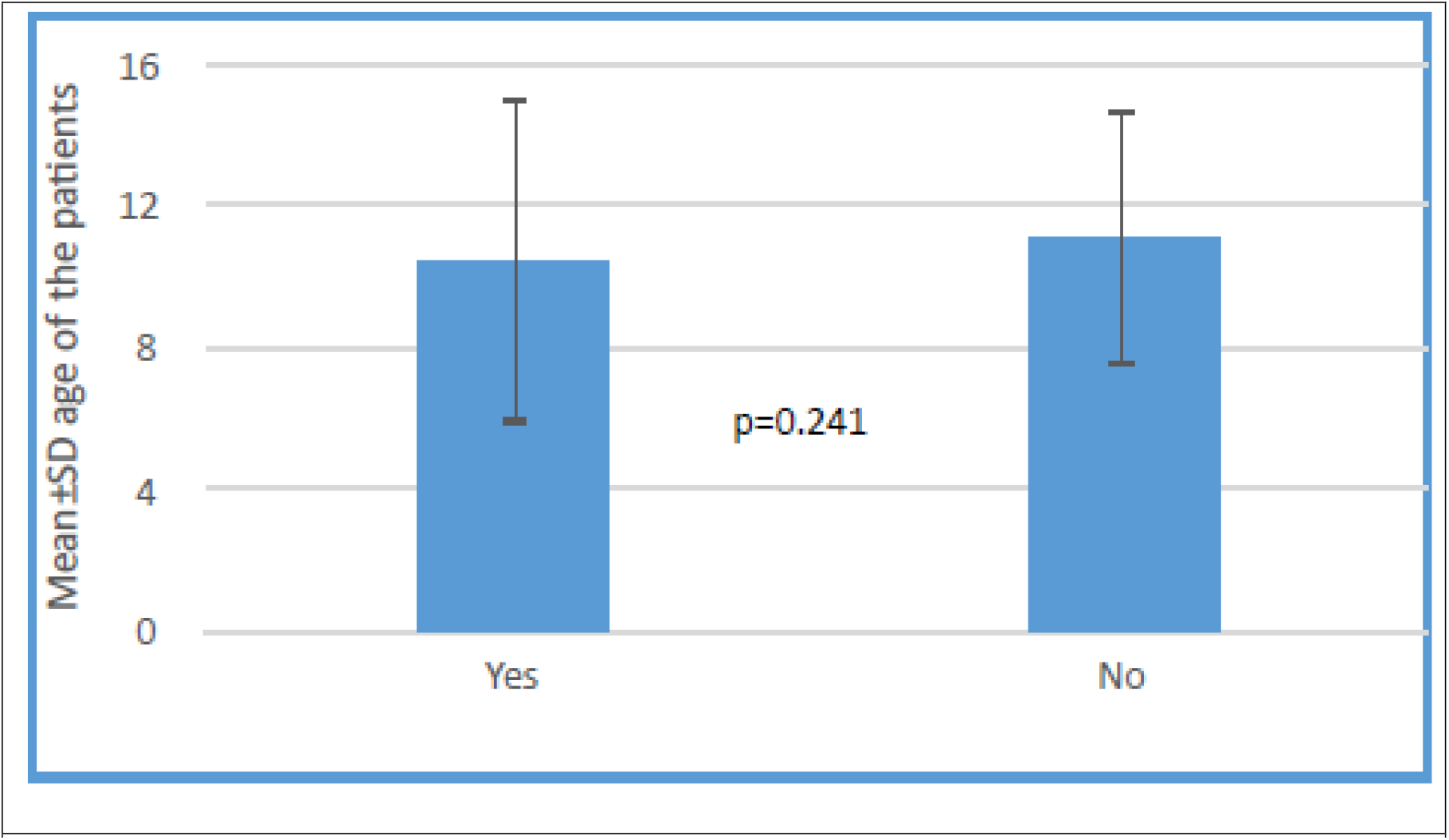
History of diabetic ketoacidosis according to age of the T1DM patients

Table 4 shows that when patients were compared according to age groups, being children (≤12 years) or adolescents 13-18 years old. Moreover, there was no statistically significant differences between children and adolescents regarding frequency of diabetic ketoacidosis and possible predisposing factors, nor the last reading of HbA1c.

**Table 4:** Differences in occurrence of ketoacidosis according to demographic characteristics of the patients

| Characteristics | History of ketoacidosis |  |  |  |  | P Value |
| --- | --- | --- | --- | --- | --- | --- |
| | Yes | | No | | $\chi^2$ | |
|  | No. | % | No. | % |  |  |
| Gender |  |  |  |  |  |  |
| • Male | 95 | 68.3 | 44 | 31.7 | 1.650 | 0.199 |
| • Female | 73 | 76.0 | 23 | 24.0 |  |  |
| Fathers' education |  |  |  |  |  |  |
| • Primary | 13 | 92.9 | 1 | 7.1 | 8.144 | 0.086 |
| • Intermediate | 28 | 68.3 | 13 | 31.7 |  |  |
| • Secondary | 58 | 76.3 | 18 | 23.7 |  |  |
| • University | 45 | 65.2 | 24 | 34.8 |  |  |
| • Postgraduate | 9 | 52.9 | 8 | 47.1 |  |  |
| Mothers' education |  |  |  |  |  |  |
| • Illiterate | 17 | 89.5 | 2 | 10.5 | 9.586 | 0.088 |
| • Primary | 17 | 81.0 | 4 | 19.0 |  |  |
| • Intermediate | 23 | 67.6 | 11 | 32.4 |  |  |
| • Secondary | 45 | 75.0 | 15 | 25.0 |  |  |
| • University | 46 | 59.7 | 31 | 40.3 |  |  |
| • Postgraduate | 4 | 80.0 | 1 | 20.0 |  |  |
| Fathers' job |  |  |  |  |  |  |
| • No job | 55 | 79.7 | 14 | 20.3 | 15.460 | 0.004* |
| • Teacher | 39 | 72.2 | 15 | 27.8 |  |  |
| • Military | 21 | 77.8 | 6 | 22.2 |  |  |
| • Physician | 5 | 62.5 | 3 | 37.5 |  |  |
| • Others | 24 | 48.0 | 26 | 52.0 |  |  |
| Mothers' job |  |  |  |  |  |  |
| • No job | 55 | 79.7 | 14 | 20.3 | 0.992 | 0.609 |
| • Teacher | 16 | 61.5 | 10 | 38.5 |  |  |
| • Others | 21 | 67.7 | 10 | 32.3 |  |  |
| Parents' consanguinity |  |  |  |  |  |  |
| • Yes, with same grandparents | 69 | 86.2 | 11 | 13.8 | 27.253 | <0.001* |
| • Yes, with different grandparents | 32 | 86.5 | 5 | 13.5 |  |  |
| • No | 61 | 55.0 | 50 | 45.0 |  |  |

\* Statistically significant

Table 5 illustrates that there was no statistically significant difference between the patients regarding history of DKA and duration since diagnosis of DM. Nevertheless, it was noted that DKA was significantly less among patients who had family history of DM than among those who did not have family history of DM (59.3% and 82.1%, respectively, p<0.001). Further analysis showed that there was no statistically significant difference within those who had familial history of diabetes mellitus according to the relation with the member who had the disease. Moreover, while the great majority of patients who had current HbA1c level above 9% (87.9%) had positive history of DKA compared to only 28.6% of patients who had their current HbA1c level ≤7%. This difference was statistically significant (p<0.001).

**Table 5:** Clinical characteristics of the patients according to their age category

| Characteristics | Age category of patients | | | | $\chi^2$ | P value |
| --- | --- | --- | --- | --- | --- | --- |
|  | No. | % | No. | % |  |  |
| History of ketoacidosis |  |  |  |  |  |  |
| • Yes | 107 | 70.9 | 61 | 72.6 | 27.4 | 0.775 |
| • No | 44 | 29.1 | 23 | 27.4 |  |  |
| Frequency of ketoacidosis |  |  |  |  |  |  |
| • Once | 51 | 47.7 | 22 | 36.7 | 4.460 | 0.347 |
| • Twice | 24 | 22.4 | 17 | 28.3 |  |  |
| • Thrice | 16 | 15.0 | 8 | 13.3 |  |  |
| • Four times | 6 | 5.6 | 8 | 13.3 |  |  |
| • Five times | 10 | 9.3 | 5 | 8.3 |  |  |
| Predisposing factors |  |  |  |  |  |  |
| • First presentation of disease | 49 | 45.8 | 21 | 35.0 | 3.021 | 0.554 |
| • Infectious cause | 7 | 6.5 | 4 | 6.7 |  |  |
| • Discontinuation of treatment | 37 | 34.6 | 26 | 43.3 |  |  |
| • Non-adherence to diet | 10 | 9.3 | 8 | 13.3 |  |  |
| • Others | 4 | 3.7 | 1 | 1.7 |  |  |
| HbA1c: |  |  |  |  |  |  |
| • $\leq 7$ | 2 | 1.9 | 4 | 6.7 | 2.480 | 0.289 |
| • $>7-9$ | 34 | 32.1 | 17 | 28.3 | | |
| • $>9$ | 70 | 66.0 | 39 | 65.0 | | |

**Table 6:** Differences in occurrence of ketoacidosis according to clinical features of the patients

| Characteristics | History of ketoacidosis |  |  |  |  | P value |
| --- | --- | --- | --- | --- | --- | --- |
|  | Yes |  | No |  | test-statistic |  |
|  | No. | % | No. | % |  |  |
| Duration of diabetes: |  |  |  |  |  |  |
| • < One year | 27 | 73.0 | 10 | 27.0 | $\chi^2=$ | 0.636 |
| • 1-5 years | 64 | 68.1 | 30 | 31.9 | 0.906 |  |
| • >5 years | 77 | 74.0 | 27 | 26.0 |  |  |
| Family history of diabetes mellitus: |  |  |  |  |  |  |
| • Yes | 67 | 59.3 | 46 | 40.7 | $\chi^2=$ | <0.001* |
| • No | 92 | 82.1 | 20 | 17.9 | 14.169 |  |
| Diabetic family members: |  |  |  |  |  |  |
| • First degree relative | 17 | 58.6 | 12 | 41.4 | $\chi^2=$ | 0.593 |
| • Second degree relative | 45 | 61.6 | 28 | 38.4 | 1.045 |  |
| • Both first & second degree relatives | 5 | 45.5 | 6 | 54.5 |  |  |
| HbA1c: |  |  |  |  |  |  |
| • $\leq 7$ | 6 | 28.6 | 15 | 71.4 | $\chi^2=$ | <0.001* |
| • 7-9 | 51 | 58.6 | 36 | 41.4 | 42.493 |  |
| • >9 | 109 | 87.9 | 15 | 12.1 |  |  |
| Age of patient (Mean±SD) | 10.5±4.6 |  | 11.1±3.6 |  | t=1.070 | 0.286 |

\* Statistically significant

## DISCUSSION

Results of this study showed that more than two thirds of the participants experienced DKA at least once. Moreover, 41.9% attributed their DKA incidents to their first presentation of the disease, while 37% attributed their incidents to discontinuation of the treatment.

This prevalence is higher than what has been reported in a Polish study, (^20)^ which reported that one-quarter of T1DM children presented with DKA at their first diagnosis, while in New-Zealand, the rate was reported to be 27% (^21)^. Nevertheless, it is lower than that reported in a Nigerian study, where about three-quarters of diabetics presented with DKA (22).

Al-Hayek et al. (^23)^, in Saudi Arabia, reported all their 103 adolescent T1DM patients had DKA, where 54.4% experienced one episode, and the main reason was the discontinuation of insulin treatment.

An American study reported stability in the rate of DKA among youth with T1DM, even though it is still high (more than third). This indicates the importance and the need for increasing awareness of signs and symptoms of diabetes and providing better access to health care. However, this rate was lower among youth with T1DM (^11)^.

Essential health education to patients and their guardians in PHCCs is essential and is considered as an effective method to decrease DKA episodes. Consequently, every consultation at a health care facility should be used ideally so that DM patients can get the maximum benefits from the health care providers. Knowledge related to “DM & DKA” must be repeated at every visit (^20)^.

The present study showed that, from the 168 DKA cases, 165 were admitted to the hospital, and only 9 reported complications with no death.

Globally, prevalence of case fatality due to DKA ranges from 0.3% to 1%, and it is mainly due to cerebral edema (^24)^.

The present study showed a significant association between DMK episodes and both consanguinity and family history.

This finding is consistent with that reported by the Satti et al. study in Al-Baha, Saudi Arabia, (^1)^ and the Zayed study, (^25)^ where consanguineous marriages ranged from 27.3% to 67.8%.

It is to be noted that, due to cultural factors, consanguineous marriage is a common practice in the Arab world, mainly first-cousins marriage. This habit is responsible for the spread of genetic diseases (^25)^.

Several studies reported that DKA is higher among female than male adolescents. This could be explained by several factors (^22;25)^. The first is attributed to puberty-associated hormonal changes, especially the raising of serum levels of some counter-regulatory hormones, e.g., estrogen, which is, by far, higher in girls than boys at puberty (^22)^. The second factor is related to body-image psychiatric problems, including eating problems, since adolescent diabetic girls often miss insulin injections for the sake of losing weight. Moreover, girls with DKA may have more behavioral problems, lower social competence, and higher levels of family struggle (^25)^.

However, in the current study, there was an association between gender and DKA episodes, but the result was not statistically significant. This is similar to the results of the Al-Hayek et al. study, where DKA was higher among females than males (^23)^.

Several studies, including our study, stated that patients with poorer glycemic control had higher risks of DKA, particularly those with HbA1c ≥10% (^25-27)^.

Results of the present study showed no significant association between age category and DKA episodes, etiology, hospitalization, and HbA1c, even though the rate was higher among children than adolescents regarding episodes, etiology, hospitalization, and HbA1c.

## CONCLUSION

More than two thirds of T1DM cases aged below 20 years, experience DKA. The main risk factors for DKA include first presentation of disease, and discontinuation of treatment. Most DKA cases become hospitalised, while less than one-tenth become complicated. Female patients are more likely to suffer from episodes of DKA than male patients. Most cases with history of DKA have uncontrolled HbA1c. DKA is higher among patients with less educated parents and among those with unemployed fathers. DKA is significantly higher among patients with consanguineously married parents.

Therefore, primary health care providers should provide the necessary health education on DKA for all T1DM patients and/or their caregivers. Health education messages should cover the main points of knowledge gaps, especially how to identify and manage sudden hyperglycaemia and raising the public’s awareness regarding DM and DKA through mass media. Patients and their caregivers should be encouraged to talk about DM with their physicians and to pay regular follow up visits for diabetes control. Further nation-wide prospective studies are needed to assess incidence of DKA and its associated risk factors.

## Data Availability

All data produced in the present study are available upon reasonable request to the authors
All data produced in the present work are contained in the manuscript

